# Pregnancy outcomes, Newborn complications and Maternal-Fetal Transmission of SARS-CoV-2 in women with COVID-19: A systematic review of 441 cases

**DOI:** 10.1101/2020.04.11.20062356

**Authors:** Rahul K Gajbhiye, Deepak N Modi, Smita D Mahale

## Abstract

**Objective:** The aim of this systematic review was to examine the maternal and fetal outcomes in pregnant women with COVID-19 and also assess the incidence of maternal-fetal transmission of SARS CO-V-2 infection.

**Data sources:** We searched PUMBED. Medline, Embase, MedRxiv and bioRxiv databases upto 3^rd^ May 2020 utilizing combinations of word variants for “coronavirus” or “COVID-19” or “severe acute respiratory syndrome” or “SARS-COV-2” and “pregnancy”. We also included data from preprint articles.

**Study eligibility criteria:** Original case reports and case series on pregnant women with diagnosis of SARS-CoV-2 infection.

**Study appraisal and synthesis methods:** We included 50 studies reporting the information on 441 pregnant women and 391 neonates. The primary outcome measures were maternal health characteristics and adverse pregnancy outcomes, neonatal outcomes and SARS-CoV-2 infection in neonates was extracted. Treatments given to pregnant women with COVID-19 were also recorded.

**Results:** Out of 441 women affected by COVID-19 in pregnancy, 387 women have delivered. There are nine maternal deaths reported. In pregnant women with COVID-19, the most common symptoms were fever (56%), cough (43%), myalgia (19%), dyspnea (18%) and diarrhea (6%). Pneumonia was diagnosed by CT scan imaging in 96 % of COVID-19 pregnant women. Pregnancy complications included delivery by cesarean section (80%), preterm labor (26%), fetal distress (8%) and premature rupture of membranes (9%). Six still births (2%) are reported. The most common co-morbidities associated with pregnant women with COVID-19 were hypertensive disorders (10%), diabetes (9%), placental disorders (2%), co-infections (3%), scarred uterus (3%) and hypothyroidism (3%). Amongst the neonates of COVID-19 mothers, preterm birth (25%), respiratory distress syndrome (8%), pneumonia (8%) were reported. There were four neonatal deaths reported. Vertical transmission rate of SARS-CoV-2 is estimated to be 8%.

**Conclusion:** In pregnant women with COVID-19, hypertensive disorders and diabetes are common comorbidities and there is a risk of preterm delivery and maternal death. Amongst the neonates born to mothers with COVID-19, respiratory distress syndrome and pneumonia are common occurrence. There are reports of still births and neonatal deaths. There is an evidence of vertical transmission of SARS-CoV-2 infection in women with COVID-19.

## INTRODUCTION

Coronaviruses are known to infect humans, other mammals, and birds causing respiratory, enteric, hepatic, and neurologic diseases ^1^. Amongst these is a novel corona virus SARS-CoV-2, first time reported from Wuhan in China in December 2019 and causes a potentially life-threating respiratory disease termed as COVID-19 ^2^. Since then, SARS-CoV-2 infection is reported from every country in the world has rapidly spread across the globe creating a massive public health problem. Owing to the high rates of human to human transmission, WHO declared COVID-19 as a pandemic ^3^. More than 3.1 million confirmed cases of COVID-19 and almost 2,24,172 reported deaths (upto 1st May, 2020), the pandemic has reportedly affected more than 180 countries/regions globally and majority of them are in local transmission phase^4^. Since this virus has not been detected in humans before, limited information is available about its effect on people; almost negligible information is available in pregnant women. Previously, members of the coronavirus family such as (SARS-CoV) and Middle East respiratory syndrome (MERSCoV) are reported to be associated with severe complications during pregnancy like miscarriage, fetal growth restriction, preterm birth and maternal deaths ^5^. During the influenza pandemic in 1918, there was a higher mortality (37%) among pregnant women as compared to mortality rate in the overall population (2.6%)^6^. During the SARS Co-V pandemic in 2003, 50% of pregnant women with SARS Co-V infection were admitted to the intensive care unit (ICU). Out of these, around 33% of pregnant required mechanical ventilation, with mortality rate of 25%^7^.

Pregnant women are particularly susceptible to respiratory pathogens and severe pneumonia, due to various factors such as physiologic changes in the immune and cardiopulmonary systems (e.g. elevation of the diaphragm, increased oxygen consumption, and edema of respiratory tract mucosa), which make them at risk of hypoxia^8^. Furthermore, during the pandemic, hospital visits may enhance the chances of infection and conversely lack of medical care during pregnancy may do more harm. Hence, there is an urgent need to devise appropriate management protocols for pregnant women to access maternal health care with minimum exposure risk is desired during the current COVID-19 outbreak. However, this would require a thorough situational analysis of COVID-19 and pregnancy.

## OBJECTIVE

The aim of this systematic review was to assess the maternal and neonatal outcomes in pregnant women with COVID-19. We also assessed if there is any evidence of the maternal-fetal transmission of SARS CO-V-2 infection. We believe this information will aid obstetricians and neonatologists to take evidence based decisions to manage pregnant women with COVID-19 and their newborns. This information will also help the societies of obstetricians and gynecologists to devise appropriate guidelines and management of pregnancy and coronavirus diseases in general.

## METHODS

### Eligibility criteria, information sources, search strategy

We performed a systematic search in PUBMED, Medline, Google Scholar, preprint servers medRxiv, bioRxiv and arXiv databases utilizing combinations of word variants for “coronavirus”, 2019 n-COV. or “COVID-19” and “pregnancy”. The time line was restricted until 3^rd^ May, 2020, no language restrictions were imposed (the articles were translated in English using google translator). We also applied snowballing method to identify any missed articles. For each search strategy, two authors (RG, DM) reviewed all the abstracts. Reviews, narrative articles, abstracts, duplicates were excluded for analysis. One article in German language could not be translated and hence excluded. Special attention was paid to exclude grey literature like media reports, blogs and information from unverified sources. Since the publication of our preprint in April 2020^9^, two studies from China were published that described clinical characteristics and outcomes of 116 and 118 pregnant women with COVID-19^10,11^. The data in these studies were directly from hospital records^10^ or from medical records extracted from National Health Commission of China^11^. We compared the information from our systematic search and those of the two studies and observed there was a considerable overlap in the patient population between the three datasets. Considering the risk of oversampling and the fact that our literature search captured information on more women from China (n=319), both these studies were excluded. The systematic review protocol was not registered due to the urgency of the matter and we did not anticipate much of evidence. Considering the nature of the studies and the outcome measures available, we could not adhere to all the guidelines of PRISMA. Although the Synthesis Without Meta-analysis (SWiM) reporting guidelines^12^ were followed. No patients or public were involved in the study design, conduct or reporting of our analysis.

### Study selection

The articles were shortlisted independently by DM and RG to include only the original studies reporting information on pregnant women with a diagnosis of SARS-CoV-2 infection (Fig. 1). The primary outcome measures were maternal clinical presentation, co-morbidities, adverse pregnancy outcomes, neonatal outcomes and SARS-CoV-2 infection in neonates. Third author (SM) coordinated the discussion for agreement of the shortlisted articles and looked for inconsistencies by randomly selecting a subset of articles (20%). The data underwent two rounds of iterations and verifications until all the inconsistencies in the data entries by RG and DM were sorted and all the authors agreed on the outcome measures. The inherent nature of the studies precluded us from ranking the quality of these studies.

**Fig 1.**
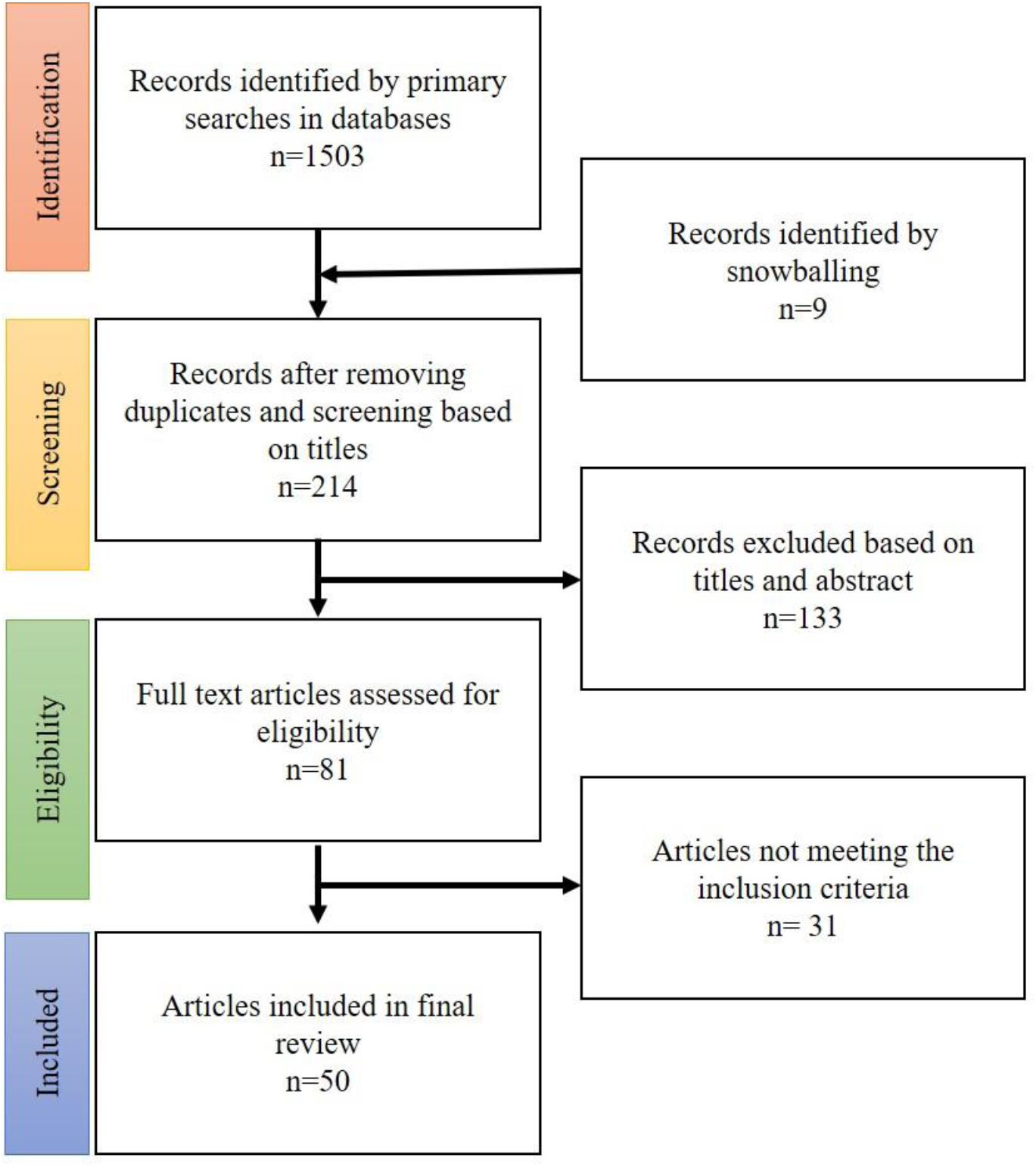
PRISMA (preferred reporting items for systematic reviews and meta-analysis) flowchart of included studies

### Data extraction

Since most were case reports and case series, individual patient data was available from these studies and entered in the table format. In the event the primary outcomes were not reported in the studies, we assumed that these did not occur in the patients and were entered as absent. Data was not available for the all the primary outcomes in all the included studies. Thus, for each primary outcome, only studies where the information was available were included for calculations and further analysis. As a secondary outcome RG independently collected data on the treatments given to pregnant women with COVID-19. As the information is sparse, it could not be organized systematically for analysis and hence it is only included in a narrative manner in the present review.

## RESULTS

### Study selection

Overall, 1503 articles were identified through database searches and snowballing. After screening and assessment of eligibility (Fig. 1); 50 studies were found eligible for inclusion and analyzed in the systematic review (Supplementary table 1). These were mainly case series and case reports from China (n=30)^10–39^, USA (n=4) ^43–46^, Iran (n=3)^47–49^, one each from Australia^50^, Canada^51^, Republic of Korea ^36^, Honduras in Central America^37^, Jordan^54^, Spain^55^, Peru^56^, Sweden^57^, Turkey^58^, Italy^59^, Portugal^60^, Switzerland^61^ and India^62^.

### Study characteristics

In most of the studies, COVID-19 diagnosis in the pregnant women was confirmed by molecular detection of SARS-CoV-2 in at least the throat swabs. Cumulatively, the data on 441 pregnant women (age range 20-49 years) was available; of which 387 have delivered which include 4 sets of twins, 4 induced abortions, 6 still births and remaining are ongoing pregnancies (Supplementary table 1). Three hundred and nineteen of the 441 pregnant women with COVID-19 are from China and the rest (n= 122) were from other parts of the world. Ninety-five percent of the women were in the 3^rd^ trimester of pregnancy and 5% of women had gestational age less than 28 weeks. Almost 50% of cases, there was a history of the women residing either in the epicenter of COVID-19 epidemic or they were in direct contact with COVID-19 confirmed cases. In the remaining women the source of infection is unknown and is possibly via a community transmission. Amongst the pregnant women with COVID-19, 80% underwent cesarean section and the rest had vaginal delivery (Supplementary table 1). The reason for cesarean section was either fetal distress or was an empirical decision made by the obstetricians in consultation with the patients. In the reported studies, cesarean sections were reported to be conducted in a negative-pressure isolation room by skillful medical team with enhanced personal protective equipments (PPEs) including N95 masks, surgical cap, double gown, double gloves, shoe covers, and powered air-purifying respirator for safe delivery.

### Maternal complications in COVID-19

The detailed breakup of the individual studies reporting maternal presentations and outcomes are given in Supplementary table 2 and the data is represented in Fig 2. The most common symptoms were fever (56%), cough (43%) and myalgia (19%). In a proportion of women dyspnea (18%) has also been reported. The major co-morbidities (Fig 2, Supplementary table 3) reported in women with COVID-19 were hypertensive disorders (10%) which included preeclampsia, gestational and chronic hypertension; diabetes (9%) including gestational diabetes, Type 1 and Type 2 Diabetes Mellitus (DM). The other co-morbidities were placental abnormalities (2%), co-infections (3%), scarred uterus (3%) and hypothyroidism (3%). Umbilical cord abnormalities were also reported in 2 cases. Placental abnormalities included placenta previa, placenta accreta and abruptio placenta. The adverse pregnancy outcomes (Fig. 2) included preterm labor (26%), fetal distress (8%), premature rupture of membranes [PROM (9%)].

**Fig 2:**
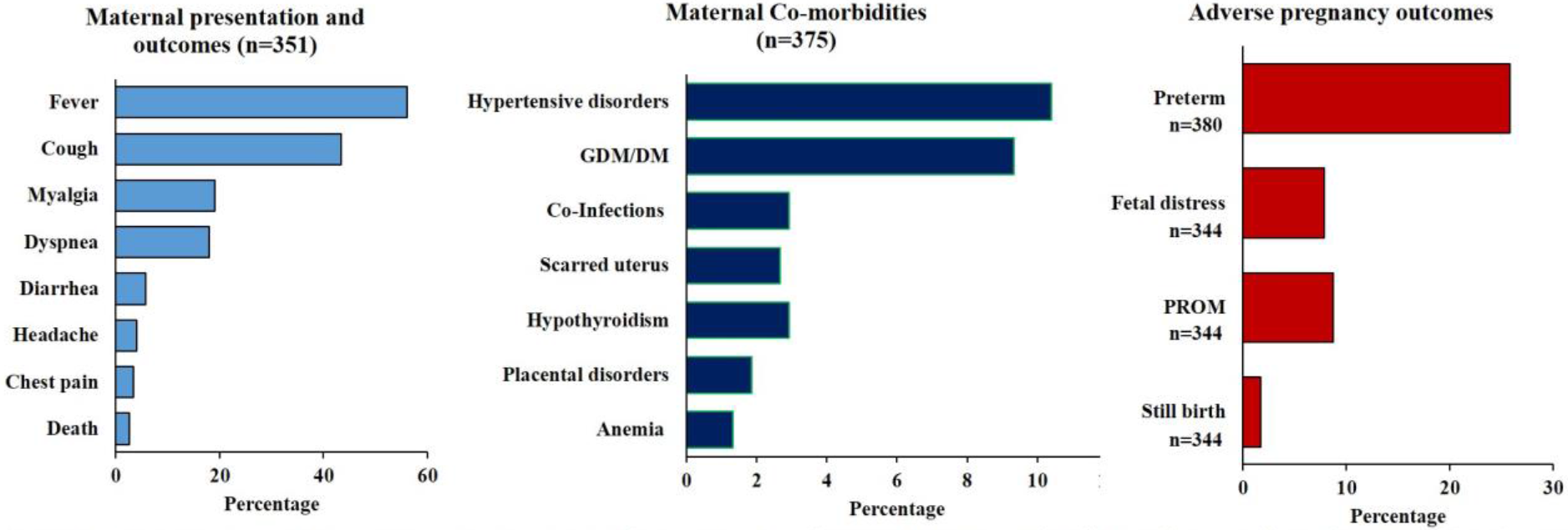
Material symptoms, co-morbidites and adverse pregnancy outcomes in pregnant women with COVID-19. Data was collated from studies reported in Supplementary Table 1. The actual values are give in supplementary table 2 and 3. n is the number of women from whom the data is derived for each outcome. GDM: Gestination Diabetes Milletus, DM: Type 1 or 2 Diabetes Milletus

Serious morbidities were reported in 11% of pregnant women with COVID-19 as they required ICU care with mechanical ventilation; of these, 10 women developed multi-organ dysfunction and were kept on extracorporeal membrane oxygenation (ECMO). Twenty four percent of women required oxygen support with nasal cannula. Due to paucity of time, we could not contact the authors of the original study to know the status of the patient kept on ECMO. Nine maternal deaths are reported amongst the studies included.

### Complications in infants born to COVID-19 mothers

Of the 391 neonates born to COVID-19 mothers, (Supplementary Table 4); neonatal data was not available from all the pregnant women with COVID-19, conversely not all maternal information was available in studies reporting neonatal outcomes of COVID-19 mothers. Thus, the data in Supplementary table 1 and Supplementary table 4 may not completely overlap.

Table 1 gives the details of the neonatal data available from these studies. In all there were 78 male and 47 female neonates (ratio 1.6) born to COVID-19 mothers. Preterm birth was reported in 25% of the neonates and Respiratory Distress Syndrome in 8%. There were six still births and four neonatal deaths reported. A proportion of neonates were admitted to the neonatal intensive care unit (NICU) with serious complications such as pneumonia (8%). Three hundred and thirteen neonates were tested for SARS-CoV-2 infections by RT-PCR and or antibody assay. In remaining neonates, the reasons for not testing were lack of reagents, non-willingness of parents and refusal to consent. SARS-CoV-2 infection was reported in 24/313 (8%) of neonates born to mothers with COVID-19. Of these, 7% were positive by RT-PCR and 3% by antibody testing method.

**Table 1:** Neonatal outcomes born to COVID-19 mothers. Data available is numbers of neonates for whom the data was reported by the authors. Nos. is numbers with the outcome and the percentage (decimal rounded off) are given.

|  | Data available | Nos. | % |
| --- | --- | --- | --- |
| Full Term | 386 | 280 | 74 |
| Preterm <37wks | 386 | 98 | 26 |
| Respiratory distress syndrome | 369 | 31 | 8 |
| Pneumonia | 369 | 30 | 8 |
| Death | 369 | 4 | 1 |
| Still birth | 369 | 4 | 1.6 |
| SARS-CoV-2 positive | 313 | 24 | 8 |

### Mother to child transmission of COVID-19

To address the extent of maternal to fetal transmission of SARS-CoV-2, we carried out a subgroup analysis where we compiled the data from the publications that explicitly reported the neonatal SARS-CoV-2 testing by the type of laboratory method used (RT-PCR or antibody or both), the neonatal samples tested and the time of testing. We further employed a strict criterion to select the studies where the diagnosis was confirmed by RT-PCR or by presence of IgM antibodies only within the first 48h of life and where the source of sampling was clearly mentioned. Table 2 gives the details of the SARS-CoV-2 infected neonates reported in the studies. In all, 261/313 neonates (84%) met the above criteria and of these, 21 tested positive for SARS-CoV-2 resulting in a possible vertical transmission rate of 8% (Supplementary Table 5). In one case amniotic fluid and in once case placenta and fetal membrane was also found to be positive for SARS-CoV-2 by RT-PCR.

**Table 2:**
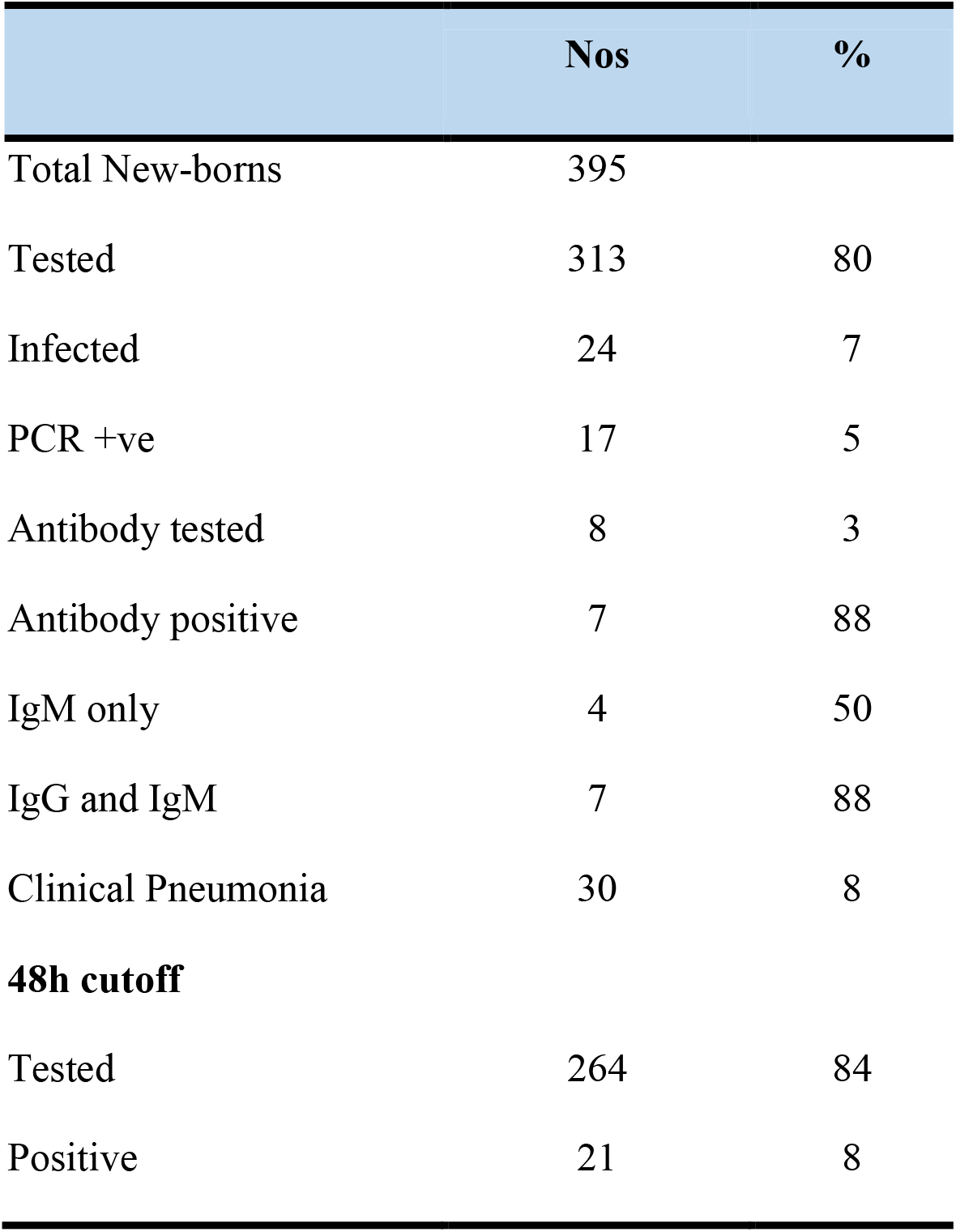
Prevalence and vertical transmission of SARS-CoV-2 from COVID-19 mothers. 48h cut-off is data is derived out of neonates tested within first 48 hours by lab test (PCR and/or antibody testing) and the sample tested is known. The percentage are rounded off to the decimal.

### Treatment and management of SARS-CoV-2 in pregnant women

Table 3 gives the treatments given to the pregnant women with COVID-19. Amongst the studies selected, 327 women were reported to receive some form of treatment for COVID-19. In the described cases and case series, most women received individual and/or combinations of several antiviral drugs (63%) and antibiotics (55%) along with the steroids mainly methylprednisolone (11%). Seven studies reported use of Hydroxychloroquine (23% of women) and four studies reported use of traditional Chinese medicine (22% of women). However, the dosages, routes of administration, duration and timings of the treatment were not detailed in most of these studies.

**Table 3:** Treatments given to mothers infected with SARS-CoV-2

|  | Nos of women | % |
| --- | --- | --- |
| Studies that reported treatment (n=33) | 327 |  |
| Antiviral (Interferons, Ganciclovir, Arbidol hydrochloride, Oseltamivir, Lopinavir, Ritonavir, Remdesivir) | 160 | 62.7 |
| Antibiotics (Azithromycin, Cefotiam hydrochloride, moxifloxacin, ceftriaxone) | 140 | 54.9 |
| Chinese herbal medicine (Jinyebaidu and Lianhuaqingwen) | 55 | 21.6 |
| Hydroxychloroquine | 59 | 23.1 |
| Steroids (methylprednisolone) | 27 | 10.6 |
| Mechanical ventilation | 31 | 12.2 |
| Oxygen with nasal cannula | 100 | 39.2 |

## COMMENT

In this large systematic review of 441 pregnant women with COVID-19 from 16 countries, we report a maternal death rate of 3%, still births (1.6%) and neonatal death rate of 1%. Hypertensive disorders and diabetes are common co-morbidities; 26% of these women delivered preterm with fetal distress and PROM being other pregnancy related complications. The adverse neonatal outcomes associated with pregnant women with COVID-19 include neonatal respiratory distress, and pneumonia. Almost these 8% of babies are born to mothers with COVID-19 had SARS-CoV-2 infection.

Previous systematic reviews and large case series on pregnant women with COVID-19 are mainly from China^10,11,63–65^. The present systematic review is generated from 50 case series and case reports from 16 countries of resulting in analysis 441 pregnant women with COVID-19. To our knowledge, this is the largest systematic review available to date generating evidence on pregnancy outcomes, complications and vertical transmission in women with COVID-19. Irrespective of the country, the clinical manifestations of COVID-19 in pregnant women were heterogeneous. Amongst the pregnant women who were SARS-CoV-2 positive, 56% presented with fever while 43% had cough. This numbers are lower than that in Chinese population which reported cough and fever in nearly 70% of pregnant women with COVID-19^11^. This observation implies that the clinical presentation of the women with SARS-CoV-2 may vary significantly in in different populations. In this regards, it is important to note that many women had mild disease and nearly 50% of the pregnant women were asymptomatic on initial presentation and were diagnosed with COVID-19 after admission for induction of labor. This was not only observed in Chinese population but also reported in women in other countries. ^32^

These results imply that asymptomatic presentations are common in pregnant women and represent a substantial risk of spreading the SARS-CoV-2 infection in the community. Given the numbers of exposed women, this is not unexpected and obstetricians must bear in mind that during the pandemic, hospitals must be prepared to deal with such atypical situations. However; the situation is alarming as it will increase the risk of exposure and infection to healthcare providers attending these women. There was an evidence of risk to the healthcare providers and four cases reported in this systematic review were physicians who acquired SARS-CoV-2 infection while providing clinical services to the COVID-19 patients ^26,29^.

Our observation highlights the need of appropriate precautions and use of protective measures especially use of personal protective equipments (PPEs) to reduce the risk of COVID-19 to the healthcare providers in obstetrics care. Our study also highlights the need of the universal screening strategy in obstetric population as many women do not present with classical symptoms or are even asymptomatic. Indeed, a recent study has shown that nearly 13% of women were afebrile but were SARS-CoV-2 positive when admitted for delivery^66^.

In this systematic review; hypertensive disorders, diabetes, and placental disorders were the top three co-morbidities identified in pregnant women with COVID-19. A nationwide study of 1590 patients with COVID-19 in China reported hypertension (16.9%) and diabetes (8.2%) as the commonest co-morbidities and risk factor for poor outcomes in COVID-19 ^34^. Beyond these, co-infections, scarred uterus, and hypothyroidism were other co-morbidities observed in the group of pregnant women with COVID-19. Currently, there is no evidence whether thyroid disease is associated with increased risk of viral infections in general and specifically COVID-19 nor is there an association between thyroid disease and severity of the viral infection.

The adverse pregnancy outcomes reported in COVID-19 women were preterm labor, fetal distress and premature rupture of membranes. For preterm births, China reports an incidence of 7.3 per 100 births or 6.7 per 100 live births^36^. However, the numbers of preterm birth observed in women with COVID-19 is comparatively higher (26%). Whether, preterm birth is a secondary complication of respiratory distress or induced directly due to viral infection needs to be determined.

Beyond preterm births, post-partum a substantial number of women required oxygen support, mechanical ventilation and ICU care. Treatment with extracorporeal membrane oxygenation (ECMO) was reported in some of the cases who became critically ill^23,46^. ECMO has been utilized in pregnancy to support oxygenation for H1N1 influenza and refractory ARDS^69^ and it is considered as an alternative rescue strategy for COVID-19^46^. Thus, post-partum women with COVID-19 are also at a risk of developing serious complications warranting emergency preparedness by obstetricians, anesthetist and pulmonologists. Therefore, it is essential that proper triage of patients should be implemented by carefully documenting prior medical and surgical history to help identifying a subset of patients who are at risk of developing serious adverse outcomes of COVID-19.

Maternal deaths due to infection are a matter of concern. The mortality rate of SARS-CoV-2 infection outside china is reported to be 1.5% and 3.6% in China ^38^. Although, there are no maternal death reported in studies from China; nine maternal deaths related to COVID-19 are reported from Iran^47,48^. The authors are aware of two maternal deaths associated with COVID-19 in India (unpublished information). It must be highlighted herein that most of the studies reported from China and elsewhere are women infected in the late third trimester or near term, the two maternal deaths reported from Iran were infected in late second trimester/early third trimester (28-30 weeks). It appears that COVID-19 in late second trimester or earlier third trimester could be more detrimental to the pregnancy. However, more data from other parts of the world on pregnant women with COVID-19 is required to get an estimate of maternal mortality in this condition.

Viral infections during pregnancy such as influenza A are reported to be associated with adverse neonatal outcomes and increased the risk of low birth weight babies^40^. Similar observation was found in pregnancy with COVID-19. Low birth weight was reported in 8% of neonates born to mothers with COVID-19 (data not shown). LBW is a risk factor for later life disease susceptibility leading to chronic morbidity. Other common neonatal complications reported are respiratory distress and pneumonia; spontaneous abortions, still births and neonatal deaths are infrequently associated with corona virus infection in general. However, there were six still births and four neonatal deaths reported. This data suggests a substantial adverse impact of maternal COVID-19 on newborns. These observations strongly suggest the requirement of special care to be given to the newborns of mothers with COVID-19.

In all, the present study estimates that 8% of the neonates born to mothers with COVID-19 have SARS-CoV-2 infection. Initial studies reported that SARS-CoV-2 was not detected in placenta, amniotic fluid, cord blood, and neonatal throat swab samples ^42, 24, 25, 44^. However, in a case series of 33 neonates born to COVID-19 women; 3 neonates were found to be positive for SARS-CoV-2 by RT-PCR^13^. Tests for IgG and IgM antibodies for SARS-CoV-2 became available in February 2020 and Dong et al., ^14^ reported a newborn with elevated IgM antibodies to SARS-CoV-2 born to a mother with COVID-19. Zeng et al., ^30^ reported IgG antibodies in 5 infants and IgM antibodies in 2 infants. However, these infants were negative for SARS-CoV-2 upon molecular testing. While the reasons for such discrepancy could be multiple, a cause of concern could be the possible false negative diagnosis by RT-PCR. It is possible that in the IgM-positive RT-PCR-negative infants the viral load may be low and beyond the sensitivity of the existing RT-PCR methods. In this context, we must highlight that 7% of neonates (even those negative for SARS-CoV-2 by RT-PCR) developed pneumonia within first two days of life. This proportion is higher than the incidence of neonatal pneumonia in general population indicating the possibility of infection by the virus and perhaps the RT-PCR has more false negatives. Therefore, further studies should include rigorous clinical assessment of the newborn along with IgM testing or employ more sensitive methods in newborn samples to determine the burden of neonatal SARS-CoV-2.

The present study aimed to answer an important question that whether SARS-CoV-2 can be transmitted from a pregnant mother to her fetus. This is more relevant based on the evidence of vertical maternal-fetal transmission of recent emerging viral infections including Zika virus, Ebola virus, Marburg virus which led to high maternal and infant mortality ^46^. Whether, SARSCoV-2 infection in the neonates was derived maternally or acquired ex utero is difficult to assess from these studies. However, we addressed this problem by applying strict criteria to include only those studies that clearly reported carrying out the diagnosis in the first 48h of life either by RT-PCR or by IgM antibodies against SARS-CoV-2. As IgM antibodies are not transferred to the fetus via the placenta ^48^, the neonates even if RT-PCR negative but positive for IgM in first 48h of life are presumed to acquire the infection in utero. The analysis revealed the possibility of intrauterine mother to child transmission, of SARS-CoV-2 in 8% of cases. The best practical approach to confirm the virus in placenta, amniotic fluid, cord blood and neonatal pharyngeal swab samples^69^. In one instance SARS-CoV-2 virus was detected in the amniotic fluid by RTPCR ^46^ one study reported viral mRNA in placental cotyledons and fetal membrane^61^. It is important to point out that in all these positive cases, the studies reported use of precautions like delivery in negative pressure rooms, wearing of N95 masks by mother and PPE by health care providers, and isolation of neonate immediately after delivery making an external acquisition unlikely. While more data is awaited, in this direction; we must consider that there is a reasonable possibility of mother to child transmission of SARS-CoV-2 and this may have long-term implications to fetal heath. Policies must be devised keeping this gap in mind towards management of COVID-19 mothers and infants.

At present, no specific treatments are available for COVID-19 and patients are symptomatically managed. In the reported studies, authors have mentioned administration of antivirals, antibiotics, steroids. Some studiers reported use of Hydroxychloroquine and some with traditional Chinese medicine in the COVID-19 pregnant women. Some of the cases included the systematic review reported incidences of thrombotic complications. Considering the hypercoagulable state of pregnancy, high-prophylactic and therapeutic dosing of anticoagulation was given in critically ill patients^49^. However, the doses administered, the time and route of administration and the length of treatment are not specified in most studies. In absence of a well reported data, it is hard to draw any conclusions on what could be the effective therapies for COVID-19 in pregnancy.

As COVID‐19 still appears to be spreading exponentially, the number of pregnant women with COVID-19 are likely to increase in different regions, countries, and continents. Therefore, at this time, it is important that all the stakeholders including pregnant women, their families, general public and healthcare providers, receive as updated, accurate and authentic information regularly. We believe that this systematic review will act as a primer for future studies and development of protocols for management of COVID-19 in pregnancy.

### Study limitations

We could not contact the authors of the original studies to find out the status of women admitted in ICU and neonates in NICU so the maternal death and neonatal death data cannot be considered updated. As there were inconsistencies observed in some reports between the results and discussion sections, we feel that there is an element of bias in the reported studies and possibility of under-reporting of the symptoms by the authors. This may influence the proportions of morbidity reported herein. Considering the nature of the studies and the urgency of the situation, we could not strictly adhere to all the criteria for PRISMA and carry out a meta-analysis.

### Conclusions

There is substantial risk of adverse pregnancy and neonatal outcomes in pregnant women with COVID-19. Evidence is garnered on preterm birth as commonest adverse neonatal outcomes; hypertensive disorders and GDM/DM and as co-morbid conditions in pregnant women with SARS-CoV-2 infection. There is an evidence of vertical transmission of SARS-CoV-2 infection in women with COVID-19. The study highlights an urgent need to bring together a multidisciplinary expertise comprising of maternal–fetal medicine and other experts globally with special emphasis on low and middle income countries to formulate evidence based clinical management guidelines for COVID-19 in pregnancy.

## Data Availability

The studies included in this systematic review are available in public domain. All data reported in the manuscript is attached and there is no additional data to be shared

## Acknowledgements

RG, DM and SM labs are funded by grants from Indian Council of Medical Research (ICMR). RG is an awardee of the DBT Wellcome India alliance clinical and public health intermediate fellowship (Grant no. IA/CPHI/18/1/503933). The manuscript bears ICMR-NIRRH ID RA/896/04–2020.

## Author Contributions

RG conceived the study. RG and DM designed the study. RG and DM screened the abstracts for inclusion in the study, extracted and analyzed the data. SM coordinated the discussion for agreement regarding potential relevance or inconsistencies and helped in data interpretation. RG and DM drafted the manuscript, which was critically revised by all authors. All authors approved the final manuscript.

## Ethical Approval

Not required

